# The Acute Treatment of Migraine with Low-Dose Naltrexone and acetaminophen Combinations and Each Component: Findings of a Small, Randomized, Double-Blind, and Placebo-Controlled Clinical Trial

**DOI:** 10.1101/2021.03.22.21254145

**Authors:** Annette C. Toledano

## Abstract

We tested two low-dose naltrexone and acetaminophen combinations and each component in the acute treatment of migraine. The patients use a single-dose of the study medication for a moderate or severe pain intensity migraine attack. Patients were adults with migraine with or without aura experiencing 2 to 20 (average 6.4) monthly migraine days. The co-primary endpoints were pain-freedom and absence of prospectively-identified most bothersome migraine-associated symptom 2 hours after dosing. We randomized 92 patients; 72 completed the study (mean age, 43 years; 75% women). Pain-freedom at 2 hours was 10.2% higher than placebo with naltrexone 2.25 mg/acetaminophen 325 mg, 10.9% with naltrexone 3.25 mg/acetaminophen 325 mg, 17.3% with naltrexone 2.25 mg, and 31.3% with acetaminophen 325 mg. The treatment groups’ migraine burden at baseline was unbalanced due to randomized patients’ uneven study completion. The acetaminophen group had the lowest migraine burden, giving its results lower credibility.

Saliently, Low-dose naltrexone alone (n=19) had a 17.3% higher response rate for headache pain-freedom at 2 hours than placebo (n=17). The naltrexone and the placebo groups were the largest and had a balanced disease burden, implying higher credibility to the naltrexone group results. We found low-dose naltrexone and acetaminophen combination, low-dose naltrexone, and acetaminophen had higher response rates than placebo in treating headache pain. The most commonly reported adverse events were sedation, nausea, and dizziness. We postulate that naltrexone’s toll-like receptor (TLR4) antagonism properties prevent pro-inflammatory cytokines’ production in the trigeminal ganglion averting “overactive nerves” (layman’s term) and migraine. Although this trial used low-dose naltrexone (defined as 1 – 5 mg/day), future phase 3 studies will test a range of naltrexone and acetaminophen combination doses.

## INTRODUCTION

The estimated global prevalence of migraine is 14.7% (that’s around 1 in 7 people).^1^ In the global burden of disease study, updated in 2013, migraine was the sixth highest cause worldwide of years lost due to disability (YLD).^2^ In the United States, approximately 38 million Americans are afflicted with migraine, and available treatments do not adequately meet the needs of many. Hence, new treatment options are needed. Oral naltrexone and acetaminophen combination, if proven effective, may provide greater efficacy/tolerability ratio than existing acute migraine treatments.

Naltrexone,^3^ an opioid antagonist approved for addiction treatment, is also an analgesic due to toll-like receptor 4 (TLR4) antagonism properties. Inhibiting the TLR4 with naltrexone in neurons of the dorsal root ganglia (DRG) and trigeminal ganglion led to reduction in pro-inflammatory cytokines’ production (calcitonin gene-related peptide [CGRP], TNF-α, and IL-1β)^5^ and reversal of neuropathic pain and migraine in animal studies.^4–10^ Naltrexone can prevent a “localized cytokine storm” (our term) in nerve cells of the trigeminal ganglion and the DRG averting pain. We postulate that naltrexone’s toll-like receptor (TLR4) antagonism properties prevent pro-inflammatory cytokines’ production in the trigeminal ganglion averting “overactive nerves” (layman’s term) and migraine.

The Interagency Pain Research Coordinating Committee (IPRCC) voted a study that used naloxone^5^ (a opioid/TLR4 antagonist similar to naltrexone) as one of 2009-2013 pain research advances that represent significant progress in the field. “This research supports TLR4 as a potential therapeutic target for treating chronic pain in patients, and, as the establishment of a completely new class of pain-relieving medication, would be a remarkable advance in pain treatment.”^11^

TLR4 is an innate (inborn) immune system receptor that usually detects invasion of foreign agents such as viruses and initiates a cascade leading to cytokines’ production to eliminate them. However, endogenous damage molecules originating from injured tissues (such as a herniated intervertebral disc) can also trigger the TLR4 leading to cytokines’ production and pain.^12^ The inborn, innate immune system is not to be confused with the learned, adaptive immune system.

Naltrexone’s analgesic properties are due to its inhibition of the production of pro-inflammatory cytokines, e.g., interleukin (IL)-6, tumor necrosis factor (TNF)-α, interferon-β, calcitonin gene-related peptide (CGRP), nitric oxide (NO), and reactive oxygen species (ROS) in nerve cells^5,8,13,14^ averting a “localized cytokine storm,” (our term) and pain generation. Naltrexone exerts its action at the beginning of the cascade leading to the production of many pro-inflammatory cytokines inhibiting their creation. Naltrexone can prevent the production of multiple cytokines eliminating the need to neutralize them after they were created.

We hypothesize that the pathophysiological event underlying migraine is the excessive production of pro-inflammatory cytokines in the trigeminal ganglion creating a neuro-inflammatory response resulting in a “localized cytokine storm.” Similarly, in the dorsal root ganglions, a “localized cytokine storm” results in neuropathic back pain. We postulate that a localized “cytokine storm” is the underlying event leading to “overactive nerves” (layman’s term) and neuropathic pain.

Pro-inflammatory cytokines exaggerate neuronal excitability, contributing to neuropathic pain and migraine. Activation of TLR4 has been implicated in the pathogenesis of migraine^9,15^ and (+)-Naltrexone blocked the development of facial allodynia in modeled migraine in rats.^9^

Dr. Bernard Bihari invented Low-Dose Naltrexone (LDN) (a daily dose of 1 to 5 mg) in the mid-1980s for “normalizing the immune system function.”^16,17^ However, scientists discovered the innate immune system and TLRs in humans in the 1990s (a Nobel Prize was awarded in 2011).^18^ The prevailing theory for LDN’s mechanism of action was that it increases endorphin production, systemically upregulating endogenous opioid signaling by a transient opioid-receptor blockade.^19^ However, later research attributed the analgesic properties of naltrexone to TLR4 antagonism leading to pro-inflammatory cytokines’ production inhibition. Currently, LDN is widely accepted as an alternative medicine modality and is used by its proponents to treat various medical conditions. It is almost sold as an everyday supplement by certain pharmacies.^19,20^

Grassroots interest in off-label LDN sprang clinical trials for fibromyalgia, multiple sclerosis, Crohn’s disease, and complex regional pain syndrome (CRPS).^21–25^

Although LDN is reportedly used as an off-label treatment for various medical conditions, there are no confirmatory studies for these off-label uses. Addiction specialists are the primary prescribers of naltrexone.

Acetaminophen potentially enhances naltrexone in several ways. Acetaminophen created synergy for analgesia in combination drugs such as Vicodin (hydrocodone/ acetaminophen).^26^ Similarly, combining naltrexone with acetaminophen could attain synergy for analgesia. Acetaminophen 1000 mg was already established as an effective acute migraine medication and can therefore enhance the treatment effect on migraine.^27,28^ Acetaminophen has the public’s trust as the world’s most consumed drug. Acetaminophen was found to be also an emotional pain reducer. Acetaminophen significantly reduced hurt feelings in human studies.^29,30^ Acetaminophen’s emotional pain reducing properties could potentially be augmented by naltrexone^31^ and enhance the combination’s effect on the overall sense of well-being.

Naltrexone’s cytokine-production inhibition properties in the trigeminal and dorsal root ganglia leading to reversal of pain could reduce pain in COVID-19 and COVID-19 vaccination. Three other (experimental) TLR4 antagonists FP7, Eritoran, and retrocyclin 101, were significantly better than placebo in treating lethal influenza.^32–34^ TLR4 signaling is a key disease pathway controlling the severity of acute lung injury.^35^ Naltrexone, a readily available TLR4 antagonist, needs to be evaluated for the treatment of pain symptoms in COVID-19.

Interestingly, the innate immune system’s pathway that mounts an initial response to viral infections is the same one that leads to neuropathic pain when triggered intrinsically by damage-molecules.

Naltrexone and acetaminophen combines naltrexone’s unutilized analgesic properties with the established and well-trusted analgesic properties of acetaminophen.

## METHODS

### Study Population

Patients were recruited to our single site in Miami, Florida, through billboards and social media advertising. The first patient was enrolled on February 14, 2017, and the last patient exited the trial on February 2, 2018.

ANODYNE-1, (the study’s name) enrolled patients 18 to 75 years of age with a history of migraine with or without aura for at least one year consistent with the diagnosis criteria of the International Classification of Headache Disorders (ICHD)-3rd edition (beta version)^36^ and have experienced between 2 and 8 migraine days in each of the 3 months before screening. Migraine onset before age 50 years was required. A history of migraine typically lasting 4 to 72 hours if untreated or treated unsuccessfully, and migraine episodes separated by at least 48 hours of headache pain-freedom were required. Patients with neurologically complicated migraine or cluster headaches were excluded. Patients using opioid medications or patients who had a history within the previous 3 years of abuse of any drug were excluded. Patients with a clinically significant hematologic, endocrine, cardiovascular, cerebrovascular, pulmonary, renal, hepatic, gastrointestinal, or neurologic disease; or had medication overuse headache to the investigator’s opinion were excluded.

Our goal was to enroll 180 patients, but we were unable to meet that goal due to hurricane Irma’s (September 2017) impact on our community.

Due to the enrollment difficulties, contrary to the original study design, we admitted patients who self-reported more than 8 migraine days per month in the previous 3 months. 18 of the 72 (25%) analyzed patients were in that category (having 9 to 20 migraine days per month and an average of 10.9). The rationale for including these patients was to evaluate the study medication’s effect on high-frequency migraine in this proof-of-concept study.

### Trial Oversight

The sponsor/investigator – Annette Toledano, M.D. was responsible for all the trial elements, including design, execution, data collection, analysis, and interpretation. The trial’s protocol and informed consent were approved by the Schulman Associates IRB (now Advarra). All patients provided written informed consent before starting the study procedures. The informed consent informed the patients of the conflict of interest of the sponsor/investigator. The patients were compensated for participation in the study. This study was conducted under an Investigational New Drug (IND) application with the United States Food and Drug Administration (FDA).

### Trial Design

ANODYNE-1 was a phase 2, single-site, randomized, double-blind, placebo-controlled, factorial, and proof-of-concept study. We evaluated the patients in two site visits; baseline/randomization and end-of-study visits. Patients treated a qualifying migraine with a single dose of the study medication within 8 weeks of the screening/ randomization visit. They returned for the end-of-study visit within seven days of using the study medication. Patients were randomized in 1:1:1:1:1 ratio to receive naltrexone 2.25 mg/acetaminophen 325 mg (n=18), naltrexone 3.25 mg/acetaminophen 325 mg (n=18), naltrexone 2.25 mg (n=20), acetaminophen 325 mg (n=18), or placebo (n=18) (Figure 1 and Figure 2).

**Figure 1:**
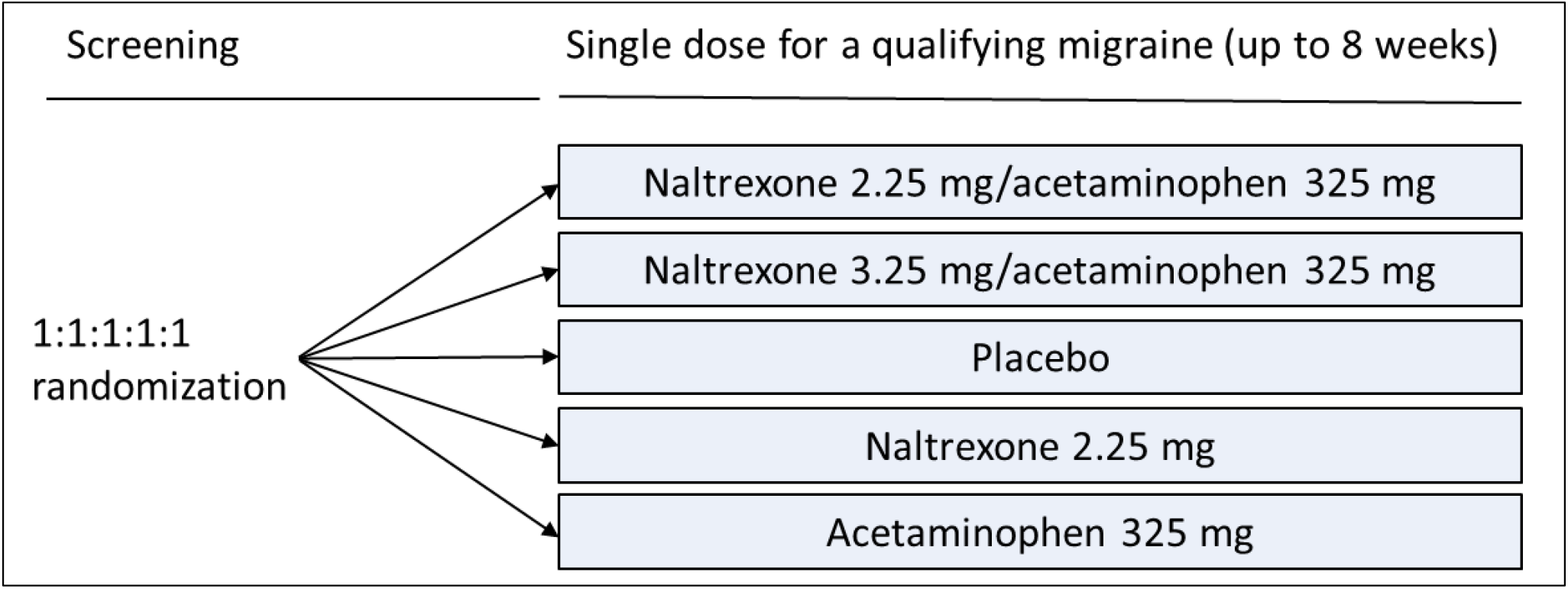
ANODYNE-1 Trial design

**Figure 2:**
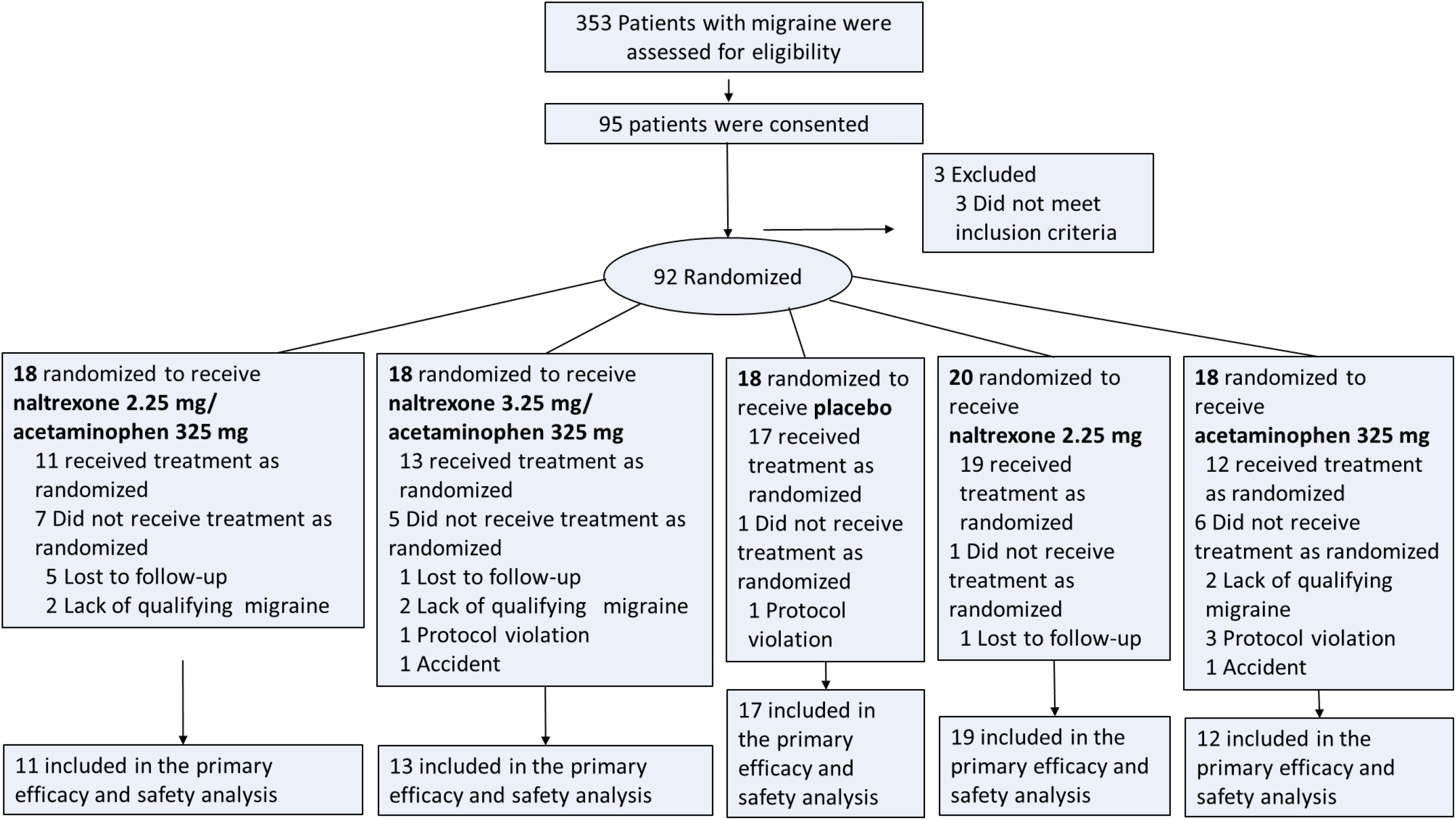
Flow chart of the ANODYNE-1, randomized trial of naltrexone and acetaminophen and its components in the acute treatment of migraine

Prospective patients completed a “migraine questionnaire” in person or by phone, and potential patients attended the screening/randomization visit. After that visit and a telephone call to confirm the blood work was adequate, the patients used the study medication to treat a qualifying migraine in the outpatient settings. Patients took one dose of study medication as soon as possible after the headache pain reached at least moderate severity. A qualifying migraine met all of the following conditions: had moderate or severe migraine headache pain, had at least one migraine headache characteristic (one-sided, throbbing, increased severity with activity); had nausea; had not taken any analgesic or migraine-specific medication in the preceding 24 hours, and have not had a headache in the previous 24 hours.

Patients were required to call the site before taking the study medication and 2 hours after to ensure they were treating a qualifying migraine and verify they completed the assessments contemporaneously. Patients did not consume rescue medications in the first 2 hours after taking the study medication. Patients used the study medication within 8 weeks from the screening visit. The final visit occurred 2 to 7 days after treating a migraine attack. Patients could take rescue medication (their usual migraine medication) beginning 2 hours after taking the study medication.

The study pharmacist prepared the study medication from marketed tablets placed in two single-ingredient capsules backfilled with microcrystalline cellulose. The study pharmacist devised the randomization schedule using a block size of 15. The medication kits were sequentially numbered, and the investigator assigned the kits to the patients consecutively.

Shortly after the study started, we required patients to have associated-nausea with their qualifying migraine as we planned to assess the study medication’s effect on nausea.

### Efficacy Assessments and Endpoint

Patients completed efficacy assessments in a paper diary. The efficacy assessments included: Headache pain severity (i.e., none, mild, moderate, or severe) and the presence or absence of migraine-associated symptoms (i.e., nausea, photophobia, and phonophobia). Patients recorded the assessments before dosing and at 7 min, 15 min, 30 min, 1, 1.5, 2, 3, 4, 6, 8, 24, and 48 hours after dosing.

The co-primary efficacy endpoints were pain-freedom and absence of prospectively-identified most bothersome migraine-associated symptom (choice of photophobia, phonophobia, and nausea) 2 hours after dosing.

Pain-freedom was defined as a reduction in headache severity from moderate or severe pain at baseline to no pain. Patients identified the most bothersome migraine-associated symptom (MBS) (choice of photophobia, phonophobia, and nausea) at the time of the qualifying migraine attack. The co-primary efficacy endpoint was the absence of what a patient had selected as the most bothersome associated symptom 2 hours after dosing.

Secondary efficacy endpoints included headache pain-freedom and MBS-absence at the other time points; sustained pain-freedom at 24 and 48 hours [(defined as having no headache pain at 2 hours after dose, with no use of rescue medication and no relapse of headache pain within 24 hours (24-hour sustained pain-free) or 48 hours (48-hour sustained pain-free) after administration of the study medication].^37^ Additional secondary endpoints included: nausea, photophobia, and phonophobia-freedom 2 hours after dosing, rescue medication used within 24 hours of treatment (defined as the proportion of patients requiring additional medication within 24 hours of dosing).

The study collected data on ‘emotional pain’ (an exploratory endpoint) for patients who reported having co-existing emotional pain (unrelated to the migraine), with the acute migraine at baseline. The emotional pain was assessed by a question in the migraine diary: “Are you currently unable to adjust or cope with a particular stress or life event causing you to experience “emotional pain” with emotions such as hurt feelings, sadness, fear, or anger?” Patients rate the emotional pain level on a 4-point rating scale at every time point.

Tolerability assessments included monitoring adverse events within 48 hours after dosing; clinical laboratory test results collected at each visit (complete blood count, hepatic and renal function), and vital signs. An electrocardiogram was obtained at the screening visit.

### Statistical Analysis

We planned for a target sample size of 36 randomized patients per treatment group (for a total of 180 patients) to provide at least 85% power to detect treatment differences between the combination and placebo based on an open-label pilot study. We were unable to attain that sample size of due to Hurricane Irma’s (September 2017) impact on our community.

The intent to treat population (ITT) included all randomized patients who received at least one dose of the study medication, recorded a baseline headache severity rating, and reported at least one post-dose assessment. The safety population included all the patients who received the study medication. The ITT Population (N=72) including 18 patients (25%) who self-reported having higher than 8 monthly migraine days at baseline.

All statistical tests were 2-sided and hypothesis tests performed at the 5% significance level. Statistical analyses were conducted using MedCalc statistical software.^38^

## RESULTS

### Patients Characteristics

We assessed 353 patients for eligibility; 92 patients met the inclusion criteria and were randomized (Figure 2). Of those randomized, 72 patients (78%) completed the study and were included in the analyzed population. The most common reason for none-completion was a loss to follow-up (8%; 7 of 92), and the second most common reason was lack of qualifying migraine (7%; 6 of 92). Patients in the analyzed population had a mean age of 43 years. Women were 75%; 85% were white; and 13% black. Eight randomized patients had pre-existing patient-doctor relationships with the sponsor/investigators (Table 1).

**Table 1:**
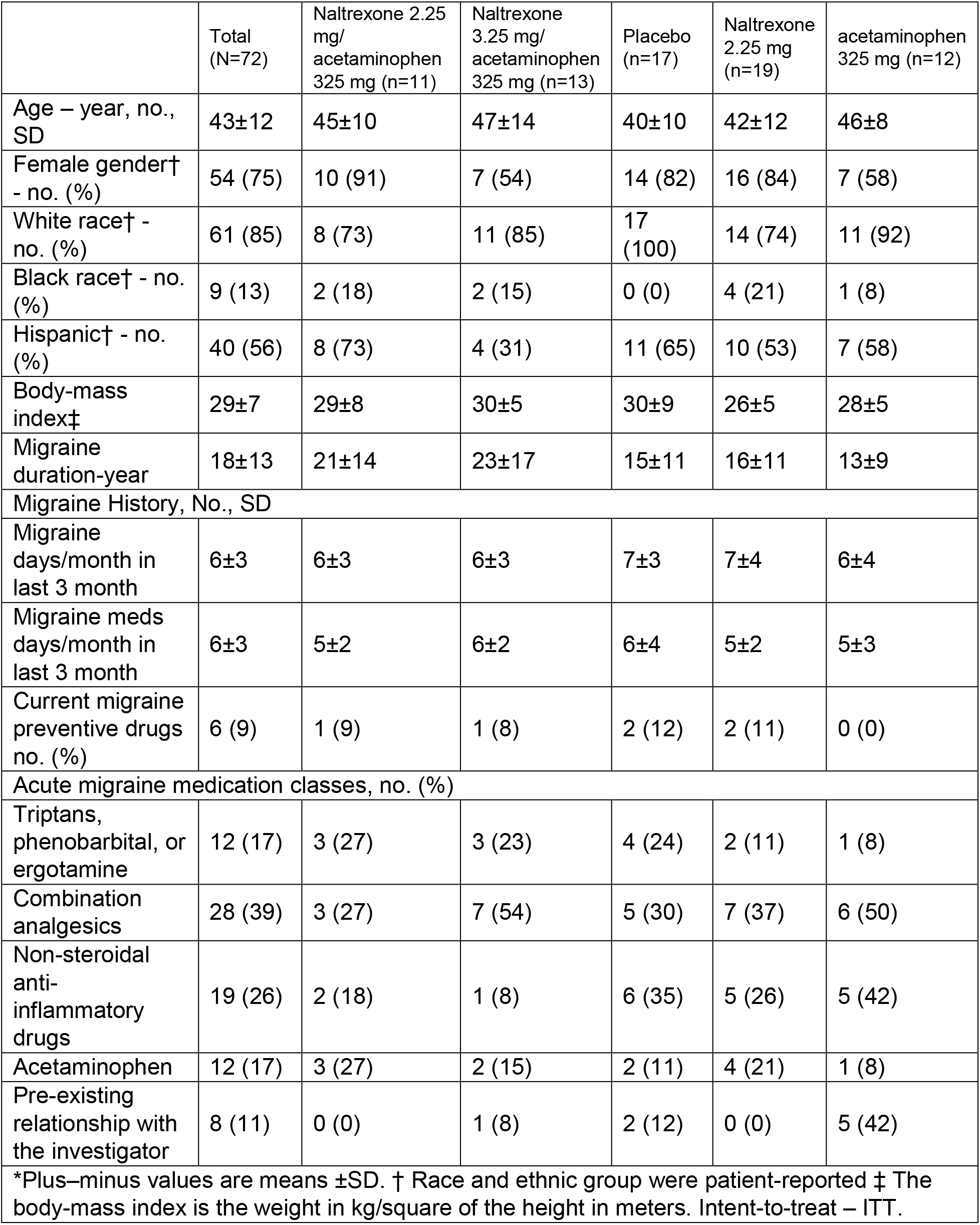
Baseline demographic (ITT Population)

In the analyzed population, 9% of patients reported current use of preventive migraine medication. Before treating a qualifying migraine attack, 54% of the patients rated their migraine headache pain as moderate, and 46% rated it as severe. At the time of the qualifying attack, 94% reported photophobia, 88% reported phonophobia, and 86% reported nausea. The most frequently reported most bothersome migraine-associated symptom was photophobia (44%), followed by nausea (33%), and phonophobia (22%). The demographic characteristics were similar among the treatment groups (Table 1).

The treatment groups’ size was markedly uneven due to varied study completion rate among the randomized patients. 7 out of 18 (39%) randomized patients in the naltrexone 2.25 mg/acetaminophen 325 mg, 6 out of 18 (33%) in the acetaminophen 325 mg, and 5 out of 18 (28%) in the naltrexone 3.25 mg/ acetaminophen 325 mg groups dropped out (Figure 2). The placebo group had 17 patients. Consequently, the results of the smaller groups, naltrexone 2.25 mg/acetaminophen 325 mg (n=11), naltrexone 3.25 mg/acetaminophen 325 mg (n=13), and acetaminophen 325 mg (n=12), were less credible. The largest group results, naltrexone 2.25 mg (n=19), which had a dropout rate of 1 out of 20 (5%), were credible.

There were baseline imbalances in clinical characteristics representing disease burden, the proportion of patients with baseline severe headache pain, associated nausea, and current use of triptans (Table 1 and Table 2). The migraine burden, measured by the proportion of patients with baseline severe headache pain, associated nausea, and current triptans use, was unbalanced between the treatment groups due to uneven completion of the study by the randomized patients and the small sample size.

**Table 2:**
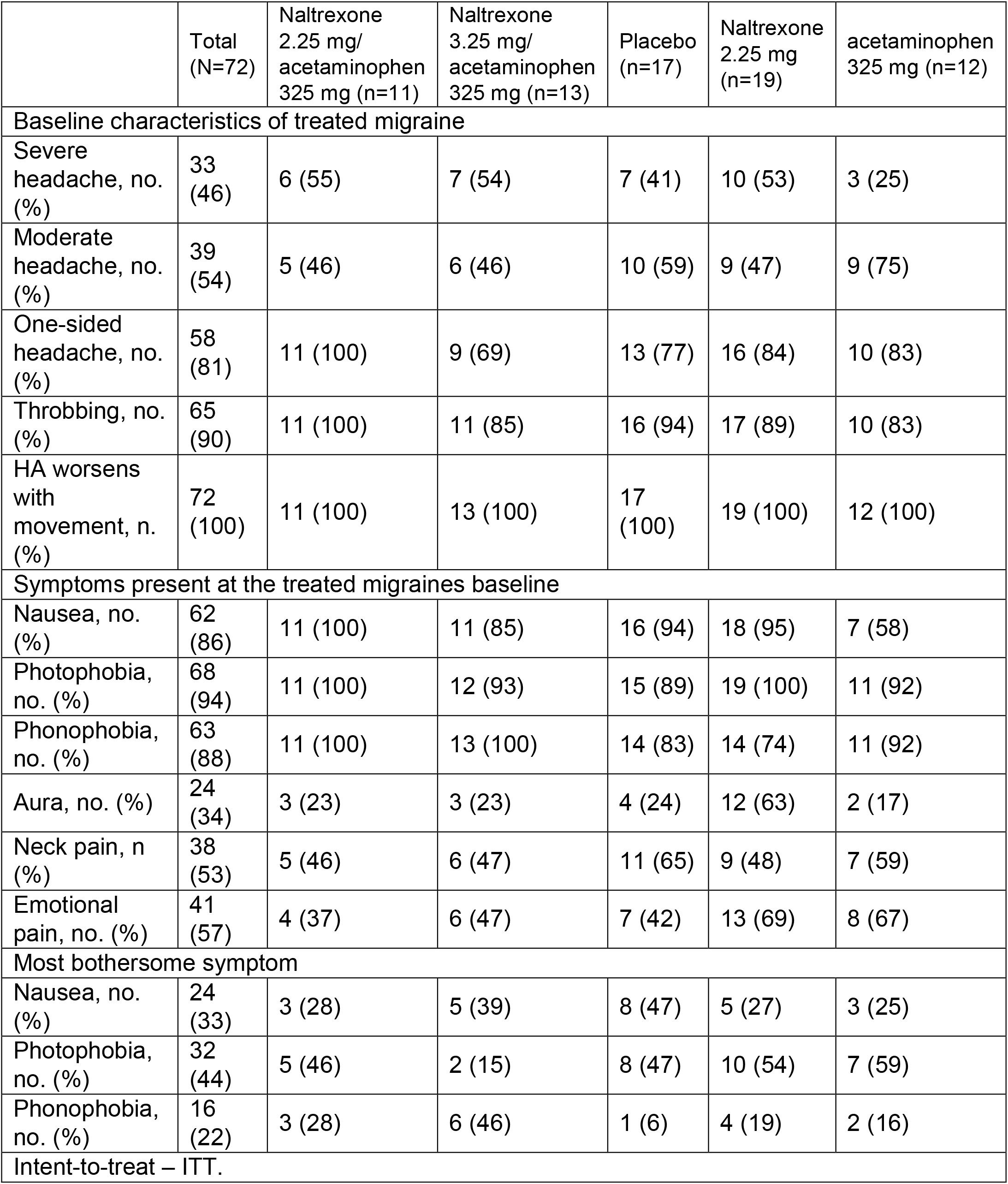
Baseline clinical characteristics (ITT Population)

The acetaminophen group had the lowest migraine burden, with a lower rate of severe headache pain (25%) versus (53-54%) in the combination and naltrexone groups, and lower associated-nausea (58%) versus (95-100%) (Table 2). The acetaminophen group had a disproportionate share of patients who had pre-existing relationships with the investigator that, in our opinion, predisposed them for the placebo effect. Five out of 8 patients with such involvement were in the acetaminophen group, constituting 42% of that group. The other groups had 0 to 12% of these patients (table 1). The patients with pre-existing relationships with the investigator had 80% pain-freedom at 2 hours regardless of the treatment assignment (much higher than the drug response rate). The higher ratio of patients with pre-existing relationships with the investigator, the lower baseline disease burden, and the small number of patients in the acetaminophen group caused an erroneously high response rate in our view. The combination groups had a higher baseline disease burden, including approximately 2-3 times higher current use of triptans than the naltrexone alone and acetaminophen alone groups, resulting in erroneously lower response rates.

The naltrexone 2.25 mg group had the most credible results since it had, together with the placebo group, the highest number of patients and a balanced disease burden.

### Efficacy Results

Pain-freedom at 2 hours was 10.2% higher than placebo in the naltrexone 2.25 mg/acetaminophen 325 mg [5 out of 11 (45.5%); P = .59], 10.9% in the naltrexone 3.25 mg/acetaminophen 325 mg [6 out of 13 (46.2%); P = .55], 17.3% in the naltrexone 2.25 mg 10 out of 19 (52.6%; P = .30), and 31.3% in the acetaminophen 325 mg [8 out of 12 (66.6%); P = .03]. The placebo group demonstrated 35.3% (6 of 17) improvement (Table 3).

**Table 3:** Efficacy Endpoints (ITT)

|  | Naltrexone<br>2.25 mg/<br>acetaminophen<br>325 mg (n=11) | Naltrexone<br>3.25 mg/<br>acetaminophen<br>325 mg (n=13) | Placebo<br>(n=17) | Naltrexone<br>2.25 mg<br>(n=19) | acetaminophen<br>325 mg( n=12) |
| --- | --- | --- | --- | --- | --- |
| Pain-freedom* |  |  |  |  |  |
| At 2-h, n (%) | 5 (45.5) | 6 (46.2) | 6 (35.3) | 10 (52.6) | 8 (66.6) |
| Diff. v. placebo, % | 10.2 | 10.9 |  | 17.3 | 31.3 |
| P-Value vs. placebo* | 0.59 | 0.55 |  | 0.30 | 0.10 |
| MBS-freedom† |  |  |  |  |  |
| 2-h, n (%) | 6 (54.5) | 8 (61.5) | 9 (52.9) | 9 (47.4) | 11 (91.7) |
| Diff. v. placebo, % | 1.6 | 8.6 |  | -5.5 | 38.8 |
| P-Value vs. placebo | 0.94 | 0.64 |  | 0.75 | 0.03 |
| Pain-freedom |  |  |  |  |  |
| At 3 h, n (%)§ | 7 (63.6) | 8 (61.5) | 7 (41.2) | 9 (47.4) | 7 (58.3) |
| Diff. v. placebo, % | 22.4 | 20.3 |  | 6.2 | 17.1 |
| P-Value vs. placebo | 0.26 | 0.28 |  |  |  |
| MBS-Freedom |  |  |  |  |  |
| At 3 h, n (%)§ | 8 (70.0) | 9 (66.7) | 10 (56.3) | 10 (52.6) | 12 (100) |
| Diff. v. placebo, % | 13.7 | 10.4 |  | -3.7 | 41.2 |
| P-Value vs. placebo | 0.49 | 0.58 |  |  |  |
| Rescue Medication |  |  |  |  |  |
| Within 24 hours, n (%) | 3 (27.3) | 5 (38.5) | 8 (47.1) | 5 (26.3) | 1 (8.3) |
| Diff. v. placebo | -19.8 | -8.6 |  | -20.8 | -38.8 |
| P-Value vs. placebo | 0.30 |  |  |  |  |
| Sustained pain-free‡ |  |  |  |  |  |
| 24 and 48 hours, n (%) | 4 (36.4) | 6 (46.2) | 6 (35.3) | 8 (42.1) | 6 (50.0) |
| Diff. v. placebo, % | 1.1 | 10.9 |  | 6.8 | 14.7 |
| Nausea-freedom |  |  |  |  |  |
| At 2 hours, n (%)² | 7 (63.6) | 7 (45.5) | 10 (56.3) | 12 (58.8) | 12 (100.0) |
| Diff. v. placebo | 7.3 | -10.8 |  | 2.5 | 43.7 |
| Photophobia-freedom |  |  |  |  |  |
| At 2-hours, n, (%)¶ | 7 (63.6) | 7 (50.0) | 11(60.0) | 10 (52.6) | 10 (81.8) |
| Diff. v. placebo, % | 3.6 | -10.0 |  | -7.4 | 21.8 |
| Phonophobia freedom |  |  |  |  |  |
| At 2-hours, n, (%)¶ | 8 (72.7) | 10 (75.0) | 12 (66.7) | 14 (64.3) | 12 (100) |
| Diff. v. placebo, % | 6.0 | 8.3 |  | -2.4 | 33.3 |
| Emotional pain score |  |  |  |  |  |
| At 2-h, sum (% of BL) | 6 (75.0) | 1 (7.1) | 7 (46.7) | 10 (41.7) | 4 (25.0) |
| At baseline, sum | 8 | 14 | 15 | 24 | 16 |
| Diff. v. placebo | 28.3 | -39.6 |  | -5.0 | -21.7 |

\*Headache pain and emotional pain score were measured on a 4-point scale (0=none, 1=mild, 2=moderate, 3=severe).
Having pain-freedom was defined as headache pain improvement from severe/moderate to none.
†The MBS, (most bothersome symptom) was measured as present=1 or absent=0. Having MBS-freedom was defined as having an absence of the pre-specified MBS, MBS=0. The MBS was prospectively identified at the treated migraine baseline as photophobia, phonophobia, or nausea.
‡24-hour sustained pain-freedom is defined as having pain-freedom 2 hours posttreatment, with no use of rescue medication and no relapse of headache pain within 24 hours. BL= baseline.
§ Adjusted for patients who took rescue medications at 2 hours, and were pain-free at 3 hours.
¶Adjusted for baseline. ITT=intent-to-treat.

The percentage of patients reporting absence of the most bothersome migraine-associated symptom at 2 hours (co-primary endpoint) was higher than placebo in the Naltrexone 3.25 mg/acetaminophen 325 mg (61.5% [8 of 13]; absolute difference, 8.6% *P* = .64) and acetaminophen 325 mg (91.7% [11 of 12]; absolute difference, 38.8% *P* = .03), but not higher in the naltrexone 2.25 mg/acetaminophen 325 mg group (54.5% [8 of 13]; absolute difference, 1.6%; *P* = .95), and naltrexone 2.25 mg group (47.4% [9 of 19]; absolute difference, −5.5%; *P* = .74). The placebo group demonstrated 52.9% (9 of 17) improvement (Table 3).

The highest response rates and separation between the combination groups and the placebo group were observed 3 hours after dosing. Treatment differences for pain-freedom rates for active drug minus placebo (Table 3) for the naltrexone 2.25 mg/ acetaminophen 325 mg were 10.2% at 2 hours and 22.4% at 3 hours after dosing. The pain-freedom rates for the naltrexone 3.25 mg/acetaminophen 325 mg dose minus placebo were 10.9% at 2 hours and 20.3% at 3 hours after dosing.

Rescue medications rate use at 24 hours were 27.3% (absolute difference, −19.8%) for naltrexone 2.25 mg/acetaminophen 325 mg; 38.5% (absolute difference, −8.6%) for naltrexone 3.25 mg/acetaminophen 325 mg; 26.3% (absolute difference, −20.8%) for naltrexone 2.25 mg, and 8.3% (absolute difference, −38.8%) for acetaminophen 325 mg. Placebo had 47.1% rescue medications use at 24 hours after dosing. Rates of rescue medication use at 48 hours were identical to the 24 hours rates.

The secondary endpoint - sustained pain-freedom from 2 to 24 hours, had response rates for naltrexone 3.25 mg/acetaminophen 325 mg, naltrexone 2.25 mg, and acetaminophen 325 mg groups higher than the placebo group.

This study did not demonstrate higher response rates for nausea in any of the naltrexone-containing groups.

The emotional pain score (hurt feelings, sadness, fear, and anger) response rate for naltrexone 3.25 mg/acetaminophen 325 mg was 39.6% higher than placebo; the naltrexone 2.25 mg, and acetaminophen were 5% and 21.7% higher than placebo, respectively. This result indicates synergy for emotional pain reduction (39.6% > 5.0%+21.7%). In contrast, the lower dose of naltrexone 2.25 mg/acetaminophen 325 mg, with the same dose of acetaminophen - 325 mg did not produce an enhanced effect in treating emotional pain, suggesting, a higher dose of naltrexone may be needed to achieve emotional pain reduction.

### Safety

Treatment-emergent adverse events were reported within 48 hours of dosing by 18% (2 of 11) of patients in the naltrexone 2.25 mg/acetaminophen 325 mg group, 31% (4 of 13) in the naltrexone 3.25 mg/acetaminophen 325 mg group, 16% (3 of 19) in the naltrexone 3.25 mg group, 25% (3 of 12) in the acetaminophen 325 mg group, and 18% (3 of 17) in the placebo group (Table 4). Patients treated with naltrexone 2.25 mg/acetaminophen 325 mg and naltrexone 2.25 mg experienced the same rate of adverse events as placebo-treated patients. Patients treated with naltrexone 3.25 mg/acetaminophen 325 mg experienced adverse events at a rate of 31% compared to 18% in patients treated with placebo. Two out of 13 patients (15.4%) in that group experienced muscle ache. The most commonly reported events were sedation, nausea, dizziness, and muscle ache. There were no serious adverse events within 48 hours after dosing. There were no deaths or discontinuations due to adverse events. There were no changes in liver function tests (Table 5).

**Table 4:** Overall summary of adverse effects by treatment group (Safety Population)

|  | Total<br>N=72 | Naltrexone<br>2.25 mg/<br>acetaminophen<br>325 mg (n=11) | Naltrexone<br>3.25 mg/<br>acetaminophen<br>325 mg n=13 | Placebo<br>n=17 | Naltrexone<br>2.25 mg<br>n=19 | acetaminophen<br>325 mg n=12 |
| --- | --- | --- | --- | --- | --- | --- |
| No. patients with any TEAE, (%) | 15 (21) | 2 (18) | 4 (31) | 3 (18) | 3 (16) | 3 (25) |
| No. patients with any serious TEAE | 0 | 0 | 0 | 0 | 0 | 0 |
| Death | 0 | 0 | 0 | 0 | 0 | 0 |
| No. patients with any TEAE related to the study medication, * (%) | 15 (21) | 2 (18) | 4 (31) | 3 (18) | 3 (16) | 3 (25) |
| Any TEAE, % of total AEs | 22 | 3 (14) | 9 (40) | 3 (14) | 3 (14) | 4 (18) |
| Sedation | 5 | 0 | 2 (15) | 2 (12) | 0 | 1 (8) |
| Nausea | 3 | 2 (18) | 0 | 0 | 1 (5) | 0 |
| Dizziness | 3 | 1 (9) | 2 (15) | 0 | 0 | 0 |
| Muscle Ache | 2 | 0 | 2 (15) | 0 | 0 | 0 |
| Dry Mouth | 1 | 0 | 0 | 1(6) | 0 | 0 |
| Dyspepsia | 1 | 0 | 1 (8) | 0 | 0 | 0 |
| Insomnia | 1 | 0 | 0 | 0 | 0 | 1 (8) |
| Nasal Congestion | 1 | 0 | 1 (8) | 0 | 0 | 0 |
| Abdominal Cramping | 1 | 0 | 0 | 0 | 0 | 1 (8) |
| Eye Twitch | 1 | 0 | 0 | 0 | 0 | 1 (8) |
| Sleepy Tongue | 1 | 0 | 1 (8) | 0 | 0 | 0 |
| Hot Flashes | 1 | 0 | 0 | 0 | 1 (5) | 0 |
| Dry Eye | 1 | 0 | 0 | 0 | 1 (5) | 0 |
| *Possibly or probably treatment-related. AE=adverse events. TEAE=Treatment emergent adverse event |  |  |  |  |  |  |

**Table 5:** Summary of any increase in hepatic function tests above the upper normal limit

|  | Naltrexone 2.25<br>mg/<br>acetaminophen<br>325 mg (n=11) | Naltrexone<br>3.25 mg/<br>acetaminophen<br>325 mg n=13 | Placebo<br>n=17 | Naltrexone<br>2.25 mg<br>n=19 | acetaminophen<br>325 mg n=12 |
| --- | --- | --- | --- | --- | --- |
| Increase in AST/GOT – no, (%) | 0 | 0 | 0 | 2 (11) | 1(8) |
| Increase ALT/GPT – no, (%) | 0 | 0 | 1 (6%) | 0 | 2 (17%) |
| Increase Total Bilirubin – no, (%) | 0 | 0 | 0 | 0 | 0 |
| Increase Alkaline Phosphatase – no, (%) | 0 | 0 | 0 | 0 | 0 |
| AST/GOT=Aspartate Aminotransferase<br>ALT/GPT=Alanine Aminotransferase |  |  |  |  |  |

### Limitations

This study had several limitations. First, the study’s sample size was too small to achieve statistical significance for the two co-primary endpoints. The small sample size resulted in baseline imbalances among the treatment groups in size and burden of disease. The study yielded credible results for the naltrexone 2.25 mg group and somewhat credible for the naltrexone 3.25 mg/acetaminophen 325 mg group. The results for the naltrexone 2.25 mg/acetaminophen 325 mg and the acetaminophen 325 mg groups were less credible.

This study had a higher-than-expected placebo response due to the sponsor-investigator status of the investigator. Specifically, patients who had pre-existing patient-doctor relationships with the investigators (8 patients) had high response rates regardless of the group assignment.

A high rate of nausea (86%) at the qualifying migraine baseline (since we required migraine-associated nausea to be present) may have delayed the study medication’s onset of action and reduced the response rate at the 2-hour time point.

This study included 18 patients, 25% of the analyzed patient, who self-reported at baseline higher than the pre-specified 2-8 migraine days per month. The patients with high-frequency migraine may have reduced response rate.

Another limitation, the exploratory endpoint of emotional pain used a direct question rather than a validated instrument to evaluate emotional pain.

This study taught us that the dose of both components of the combination needs to be assessed in future studies. Using a dose that was too-low was a potential flow of this study. We plan future studies to be sponsored, adequately powered, and not require baseline nausea. We also plan to verify in a run-in period that the patients are within the pre-specified monthly migraine days.

## DISCUSSION

In this study, naltrexone 3.25 mg/acetaminophen 325 mg demonstrated higher response rates than placebo for both co-primary endpoints. Naltrexone 2.25 mg/acetaminophen 325 mg demonstrated higher response rates than placebo only for the pain-freedom co-primary endpoint, suggesting the 2-hour MBS-absence may requires the combination. Naltrexone 2.25 mg achieved higher response rates than placebo for the 2-hour pain-freedom but not for the 2-hour MBS-absence, suggesting naltrexone may primarily effect migraine pain but not MBS.

The combination containing the higher dose of naltrexone had higher response rates in multiple endpoints suggesting a dose-response for the naltrexone component.

This study demonstrated response rate synergy of the naltrexone 3.25 mg/ acetaminophen 325 mg over the individual components for emotional pain reduction.

The higher responses at the 3-hour time point than the 2-hour time point with the combination groups for both co-primary endpoints suggest a benefit of the combination relative to placebo. The 3-hour endpoint results may have occurred sooner if not for double the prevalence of nausea relative to other acute migraine clinical trials. Nausea may have delayed the absorption of the study medication.

Naltrexone 2.25 mg had a 17.3% higher than the placebo response rate for the 2-hour pain freedom. The naltrexone 2.25 mg and the placebo groups had the highest number of patients, 19 and 17, respectively, making these results, although not statistically significant, credible. Recently approved oral acute migraine medications had 7.6% to 16.9% higher than placebo response rates.

The response rate for acetaminophen 325 mg for the 2-hour pain freedom over placebo was 31.3%. We believe this response rate was erroneously high due to a low disease burden and three times the rate of patients with pre-existing relationships with the investigator.

The combination groups had a higher baseline disease burden, as reflected by several measures. The acetaminophen alone and naltrexone alone groups had higher response rates than the combination groups (at either dose). In a study with balanced baseline burden, the combination’s response is expected to be at least as high as one component alone. MBS-absence 2 hours after dosing for naltrexone 2.25 mg over placebo was −6%, indicating possible lack of efficacy of naltrexone alone for MBS. This study failed to demonstrate a response for nausea for any of the naltrexone-containing treatment groups.

The response rate for sustained pain-freedom from 2-24 hours for naltrexone 3.25 mg/acetaminophen 325 mg was higher than placebo by 10.9%; naltrexone 2.25 mg was 6.5% higher. Naltrexone 3.25/acetaminophen 325 mg was higher than naltrexone by 4%. This endpoint is recommended by the FDA for the combination rule requirement.

All the study treatments were well-tolerated. Adverse effects included mild and transient sedation, nausea, dizziness, and muscle ache.

## CONCLUSIONS AND FUTURE STUDIES

Given the higher response rates for headache pain and emotional pain in the combination with the higher naltrexone dose, our future studies will test a range of naltrexone and acetaminophen combination doses.

It would be prudent to develop a rapid-acting formulation to achieve a therapeutic effect within 2 hours after dosing (an FDA’s requirement for an acute migraine drug).^37^

This study is first to demonstrate response rates higher than placebo for naltrexone in the acute treatment of migraine. This study used approximately 1/20 of approved naltrexone’s tablet. Targeting the toll-like receptor-4 would represent a novel approach to treating migraine. Adding acetaminophen, the world’s most consumed drug, could enhance the combination’s analgesic effect and boost the public’s trust conferring an advantage over naltrexone alone. This trial provides preliminary evidence for the potential benefits of naltrexone as a pain reducer. Adequately powered clinical trials are needed to confirm this study’s findings. The study’s sample size was too small to achieve statistical significance for the two co-primary endpoints.

## Data Availability

Data not available

## PATENTS

The naltrexone and acetaminophen combination received two U.S. patents for treating pain and a U.S. patent for treating emotional pain. The emotional pain patent was based on this study’s naltrexone 3.25 mg/acetaminophen 3.25 mg synergism data.

### Author Contribution: Concept, design, Statistical analysis, and data interpretation

Annette Toledano.

Dr. Toledano had full access to all of the data in the study and takes responsibility for the integrity of the data and the accuracy of the data analysis.

### Study Pharmacist

Ayman Mohamed PharmD.

### Funding/Support

This trial was fully funded by Allodynic Therapeutics, LLC.

### Grants

None

### Trial Registration

ClinicalTrials.gov Identifier: NCT03061734

### Institutinal Review Board (IRB)

This trial’s protocol and informed consent were approved by the Schulman Associates IRB (now Advarra).

### Investigational New Drug Application

This study was conducted under an Investigational New Drug (IND) application with the United States Food and Drug Administration (FDA).

## Acknowledgments

We acknowledge the immense contribution of the study patients, the study pharmacist, and the clinical research coordinators, whose hard work made this trial possible.

